# A multi-view similarity network fusion framework for syndrome discovery from aggregated health records

**DOI:** 10.64898/2026.06.30.26356975

**Authors:** Ana Paula Gomes Ferreira, Aleksandar Anžel, Pilar Tavares Veras, Pablo Ivan Pereira Ramos, Manoel Barral-Netto, Izabel Marcilio, Georges Hattab

## Abstract

Syndrome discovery, the identification of clinically meaningful groupings of signs and symptoms, is a foundational but labor-intensive task in syndromic surveillance, and the COVID-19 pandemic exposed the rigidity of expert-curated definitions in the face of novel threats. Unsupervised, data-driven methods are well-suited to this problem but remain underused. We propose an unsupervised framework based on Similarity Network Fusion (SNF) that operates on only five variables: diagnosis code, sex, age group, epidemiological week and year, and encounter count. Each diagnosis code was represented through three complementary views corresponding to the fundamental questions of syndromic surveillance: what condition is recorded (clinical, via SapBERT embeddings), who is affected (demographic, via chi-square distances), and when it occurs (temporal, via Move-Split-Merge). The fused affinity matrix is partitioned by spectral clustering and exported directly in the Open Syndrome Definition (OSD) format for downstream integration. To our knowledge, this is the first framework of SNF applied to the task of syndrome discovery. We use the framework in 72.9 million primary care encounters across ten Brazilian municipalities. Treating each city as an experiment with no shared training signal yields 47 candidate syndromes, 72% of which are rated fully valid by an expert blind to the procedure. By requiring no predefined targets, the framework discovers candidate syndromes at scale, including ones never explicitly sought, and emits them in a deployable format, shortening the path from emerging signal to usable definition.

**Author summary:** Public health systems are under growing pressure to catch outbreaks early, but the syndrome definitions that trigger surveillance alerts, the combinations of signs and symptoms that flag a possible threat, are still built largely by hand and too slowly to keep up. We developed a data-driven framework that groups diagnosis codes into candidate syndromes by asking three questions: what condition a code represents, who is affected, and when it occurs. Applied to 72.9 million primary care encounters across ten Brazilian cities, the framework recovered known syndromes, including arboviruses and influenza-like illness, without being given their definitions, and produced 47 candidate syndromes that held across all cities, 72% of which a domain expert rated valid. Beyond infectious disease, it surfaced coherent groupings in areas that surveillance rarely monitors, such as mental health. Each candidate is exported in the Open Syndrome format, ready for existing surveillance systems and the wider community to use.

## Introduction

Syndromic surveillance systems monitor health-related data in near real time to enable the early detection of outbreaks and other health threats before traditional surveillance reports are available [1, 2]. These systems depend on syndrome definitions (also called syndromic indicators [1]): combinations of symptoms, chief complaints or clinical diagnosis extracted from patient records and used to represent a public health threat [3]. In practice, syndrome definitions are created manually by domain experts, validated by epidemiological review, and integrated into the surveillance infrastructure. It works for well-characterized conditions such as Influenza-Like Illness (ILI) or acute diarrheal disease, but is slow, labor-intensive, and rigid [4, 5]. Updating or creating new definitions can take months, and the resulting definitions may not reflect local epidemiological patterns in geographically heterogeneous territories [6].

The limitations of predefined syndromes became evident when existing definitions failed to capture the early clinical presentation of a novel pathogen during the COVID-19 pandemic [7], or when the Zika virus circulated undetected in Latin American countries for nearly one year before it was officially reported [8]. These experiences highlighted the importance of syndrome discovery: the automated identification of clinically meaningful groups from health data, without requiring prior definitions as input. A recent scoping review found that only three of fifteen publications on AI-assisted syndrome discovery targeted non-specific syndromes, and the majority relied on supervised methods applied to English-language free-text clinical notes [5]. Unsupervised approaches capable of operating on structured, aggregated, privacy-compliant data remain scarce.

Processing free-text notes carries privacy and operational costs that scale poorly with routine, real-time surveillance, whereas structured administrative data are already collected for reporting, easier to govern, and shareable in aggregated form. Structured, aggregated data are thus a natural substrate for syndrome discovery at scale.

In this work, we propose a general unsupervised framework for syndrome discovery that operates on five minimal variables available in most administrative health databases: diagnosis code, sex, age group, time index, and encounter count. A diagnosis code carries clinical, demographic, and temporal signals of different geometries, so no single representation captures it; the framework models each code through three complementary views that correspond to the fundamental questions of syndromic surveillance [3, 9]: what condition is being recorded (clinical view, via biomedical language model embeddings), who is affected (demographic view, via sex and age distributions), and when it occurs (temporal view, via chronological time series alignment). These heterogeneous representations are integrated using Similarity Network Fusion (SNF) [10], which iteratively reinforces cross-view agreement while attenuating view-specific noise. To our knowledge, this is the first application of SNF to syndrome discovery.

As a case study, we instantiate the framework on 72,936,819 primary care encounters spanning ten Brazilian municipalities across the five macro-regions, treating each city as a self-contained experiment with no shared training signal. In many settings, primary care remains an underused surveillance source [11–13] that records conditions before they reach hospital settings, and the Brazilian records encode diagnoses across three coding terminologies (ICD-10, ICPC-2, and a local administrative code), a heterogeneity the framework resolves through a shared embedding space rather than manual crosswalks.

The main contributions of this work are:

- **A multi-view SNF framework for syndrome discovery.** The three-view design (clinical, demographic, temporal) maps directly onto the *what*, *who*, and *when* questions that define syndromic surveillance, and operates on five common collected variables without requiring patient-level data, free-text notes, or specialized hardware.
- **Geographic replication as an evidence standard.** Running the pipeline independently on multiple sites with no shared training signal lets cross-site consistency act as external validation that needs neither labeled data nor predefined definitions. A syndrome that recurs across sites with distinct epidemiological profiles and coding practices is unlikely to be a local artifact.
- **An anchor-calibrated consistency threshold.** The pairwise consistency criterion is calibrated against a known epidemiological reference rather than set arbitrarily, a contribution applicable to any multi-corpus clustering evaluation where a ground-truth anchor exists.
- **The aggregated recall metric.** A practical adaptation of standard recall that credits clinically meaningful fragmentation of reference syndromes into coherent sub-syndromes, addressing a situation that arises naturally in unsupervised discovery but is penalized by conventional metrics.
- **Direct operational integration via the Open Syndrome Definition (OSD) format.** Each candidate syndrome is exported in the OSD format [14], reducing the cycle from discovery to usage and addressing one of the core bottlenecks identified in the syndrome definition literature [5].
- **An empirical case study at national scale.** Application to 72,936,819 primary care encounters across ten Brazilian municipalities recovers two published reference syndromes without their definitions provided as input, and yields 47 candidate syndromes of which 72 % are rated fully valid by a domain expert blind to the clustering procedure. The full implementation is publicly available at https://github.com/anapaulagomes/virola.

## Methods

### Data source and data cleaning

To demonstrate the framework, we conducted a retrospective case study using aggregated routinely collected data from the Brazilian national Primary Health Care (PHC) database. Brazil is an upper-middle-income country with approximately 212.6 million inhabitants across 5,570 municipalities [15]. The Brazilian Unified Health System (SUS) is one of the largest public health systems globally, providing comprehensive, free-at-the-point-of-delivery care to the entire population [16]. Managed by the Ministry of Health (MoH), the PHC database is a hierarchical, decentralized information system that compiles data on all PHC encounters to support resource allocation from the federal to the municipal level.

We obtained the aggregated data with permission from the MoH. All PHC records are coded based on the chief complaints, and the source suppressed the rarest 2% of diagnosis codes to prevent re-identification.

The dataset comprises 72,936,819 primary care encounters, aggregated by diagnosis code, sex, age group and date and city of encounter into 17,174,982 rows across ten municipalities in Brazil’s five macro-regions and two population brackets (250,000–500,000 and *>*500,000 inhabitants), spanning the full years 2022 through 2024 (see Fig 2 and Table 1). Unlike hospital datasets, which typically adopt a single coding system, primary care data in Brazil are heterogeneous, encoding diagnoses and conditions simultaneously across three distinct medical terminologies:

**Table 1.**
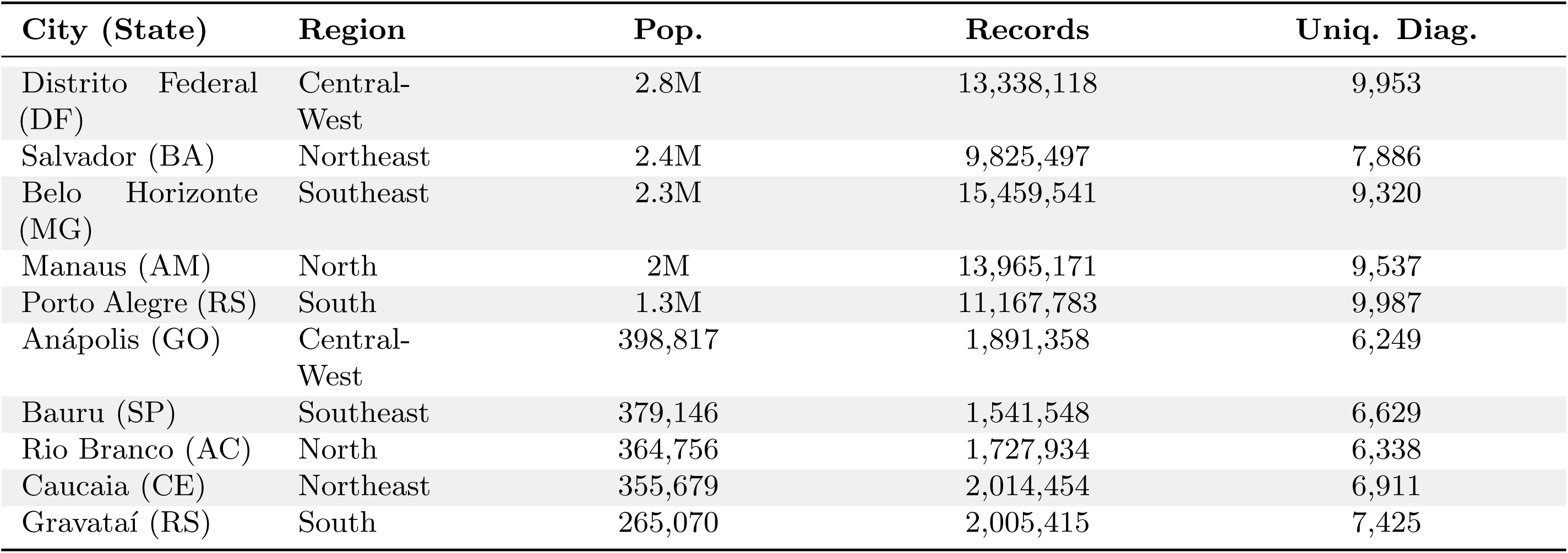
City demographics and data overview. . This table lists the cities included in this study, along with their regional characteristics, estimated population according to the Instituto Brasileiro de Geografia e Estatística (IBGE) [15], and statistics on the number of records generated from 2022 to 2024.

| City (State) | Region | Pop. | Records | Uniq. Diag. |
| --- | --- | --- | --- | --- |
| Distrito Federal (DF) | Central-West | 2.8M | 13,338,118 | 9,953 |
| Salvador (BA) | Northeast | 2.4M | 9,825,497 | 7,886 |
| Belo Horizonte (MG) | Southeast | 2.3M | 15,459,541 | 9,320 |
| Manaus (AM) | North | 2M | 13,965,171 | 9,537 |
| Porto Alegre (RS) | South | 1.3M | 11,167,783 | 9,987 |
| Anápolis (GO) | Central-West | 398,817 | 1,891,358 | 6,249 |
| Bauru (SP) | Southeast | 379,146 | 1,541,548 | 6,629 |
| Rio Branco (AC) | North | 364,756 | 1,727,934 | 6,338 |
| Caucaia (CE) | Northeast | 355,679 | 2,014,454 | 6,911 |
| Gravataí (RS) | South | 265,070 | 2,005,415 | 7,425 |

**ICD-10** International Classification of Diseases, 10th Revision [17] (abbreviated as CID-10 in Portuguese): an alphanumeric coding system maintained by the World Health Organization (WHO), it includes 14,446 codes (11,432 represented in the dataset). It is used to categorize medical diagnoses, symptoms, and procedures, with a focus on the disease.

**ICPC-2** International Classification of Primary Care, 2nd Edition [18] (abbreviated as CIAP-2 in Portuguese): it prioritizes the reason for the health encounter (signs, symptoms, and complaints), and it is the standard coding system in the Brazilian PHC. It includes 686 codes (all of which are represented in the dataset).

**AB** the Portuguese abbreviation for *Atenção Básica* (Primary Care) code [19]: a legacy Brazilian chief complaints coding integrated into the national digitization strategy [20]. It remains in active use in low-connectivity settings where records are collected on paper and digitized asynchronously. It includes 23 codes (22 represented in the dataset).

Each terminology encodes a complementary clinical perspective: ICD-10 for disease classification, ICPC-2 for encounter-driven documentation, AB for conditions managed in resource-constrained settings. Their coexistence reflects the structural complexity of Brazil’s Universal Health System.

Raw records contains with a combined CIDCIAP field encoding both terminology type and code (*e.g.* ICD(J06), CIAP2(R74), AB(ABP006)); parsing separated these into code type and code. We discarded records missing year, epidemiological week, or age group (see S2 Fig and S3 Fig). We enriched records with city name, region, and population range via Instituto Brasileiro de Geografia e Estatística (IBGE) municipality codes [15]. We randomly selected ten cities with populations of at least 250,000 inhabitants, targeting macro-regional hubs with sufficient data volume for stable distance estimation.

Before applying the SNF framework for syndrome discovery, a domain expert manually labeled the diagnostic codes accounting for the most frequent 90% of aggregated encounter-level data in the dataset (see S4 Fig). All codes describing specified signs, symptoms, or diagnoses of diseases or health conditions were labeled as relevant. Codes corresponding to life stages (*e.g.*, pregnancy, menopause), behavioral factors (*e.g.*, smoking), healthcare utilization and administrative encounters (*e.g.*, preventive medicine/health maintenance), or acute trauma and sequelae were classified as non-indicative and were excluded from the analysis (see examples in Table 2). We retained all codes not assessed in this phase. The labeled list is available as supplementary material S1 Table.

**Table 2.** Examples of non-indicative codes. . The table shows examples of codes labeled as non-indicative from the three medical terminologies (ICD-10, ICPC-2, and AB) used in this work.

| Code | Code Type | Description |
| --- | --- | --- |
| A98 | ICPC-2 | Preventive medicine/health maintenance |
| ABP003 | AB | Sexual and reproductive health |
| R68 | ICD-10 | Other general symptoms and signs |

Additionally, we removed ICD-10 codes belonging to the Z chapter (factors influencing health status and contact with health services), as they encode administrative encounters rather than clinical diagnoses (*e.g.* Z76.0, issue of repeat prescription). We also excluded codes observed in fewer than 12 distinct epidemiological week-year pairs, ensuring that temporal series were long enough to reflect a seasonal or epidemic pattern rather than noise. To reduce sparsity in the demographic view, we aggregated the original age strata into four epidemiologically significant bins: Infants (*<* 1 year), Children/Adolescents (1–15 years), Adults (16–59 years), and Elderly (*>* 60 years).

#### Research ethics

The study is based on secondary, aggregated, non-identified data and was approved by the Ethical Review Board of the Oswaldo Cruz Foundation–Instituto Gonçalo Moniz, Fiocruz Bahia, CAAE 61444122.0.0000.0040. All study procedures followed the ethical standards for research involving human beings, defined by resolution 466/2012 of the Brazilian National Health Council. The ethics committee granted a waiver for the need to obtain informed consent for data collection as the study was based on an aggregated database consisting of the number of encounters, per epidemiological week, per municipality, and per diagnostic code, with no information at the individual level.

### Similarity Network Fusion

Each diagnosis code in the dataset can be characterized along three complementary dimensions: its clinical meaning, the demographic profile of the patients who receive it, and the temporal pattern of its occurrence. These dimensions are heterogeneous, spanning unstructured text, count-based compositional vectors, and ordered time series; no single distance measure is appropriate for all three. Multiview learning addresses this by building a separate similarity structure for each dimension and integrating them into a representation richer than any single view.

Similarity Network Fusion, introduced by Wang *et al.* [10], constructs one affinity network per view, where nodes are items (here, diagnosis codes) and edge weights reflect pairwise similarity under that view’s metric. Fusion proceeds through an iterative non-linear diffusion process in which each network is updated by propagating information from the others, reinforcing edges consistent across views while attenuating view-specific noise. The fused matrix captures structure shared or complementary across views and has been shown to outperform single-view representations for clustering in heterogeneous biomedical data [10].

Originally demonstrated on genomic data, SNF has since been applied across biomedical domains, including cancer subtype discovery [21], patient stratification from multi-omics profiles [22], and disease phenotyping from electronic health records [23].

The framework applies SNF over three views constructed from the five available variables. The choice of three views follows the three questions a syndromic surveillance system must answer [3, 9]: what clinical condition is recorded, who is affected, and when it occurs. These map onto the clinical, demographic, and temporal views described next.

For each view, the pairwise distance matrix is first normalized to [0, 1] to place all three views on a comparable scale. An affinity matrix is then derived using an adaptive Gaussian kernel with local scaling [10]: the kernel bandwidth for each node pair is estimated from the mean distance to its 20 nearest neighbors, scaled by a factor *µ* = 0.5. Each affinity matrix *W* is then rescaled to [0, 1] by zeroing the diagonal and dividing by max(*W*). Because the adaptive kernel produces affinities whose magnitude depends on each view’s local density, this rescaling prevents any single view from dominating the fusion through its kernel scale alone. The three affinity matrices are fused using the snf library [24], which implements the diffusion procedure of Wang *et al.* with *k*_fuse_ = 10 neighbors defining the local neighborhood during diffusion. Affinity construction uses 20 nearest neighbors for local scale estimation, while *k*_fuse_ sets the neighborhood for diffusion.

#### View construction

Each diagnosis code is represented in three views capturing complementary aspects of its surveillance-relevant behavior (Table 3). Within each view, pairwise distances are computed with a metric matched to that representation’s geometry, and the resulting matrices feed the SNF pipeline.

**Table 3.** Summary of the three views used in the SNF pipeline. Each view captures a distinct surveillance-relevant dimension of a diagnosis code, characterized by a dedicated representation and distance metric chosen to match the geometry of the data.

| View | Question | Representation | Distance |
| --- | --- | --- | --- |
| Clinical (*what*) | What does it represent? | SapBERT embeddings; ICD-10, ICPC-2, and AB codes unified in a shared embedding space | Cosine |
| Demographic (*who*) | Who is affected? | Diagnosis × sex/age-group bin matrix, row-normalized to probability distributions | Chi-square |
| Temporal (*when*) | When does it occur? | Diagnosis × chronological epidemiological week-year series, row-normalized to $[0, 1]$ | Move-Split-Merge (MSM) |

**Clinical view.** The clinical view encodes the semantic meaning of each diagnosis code as a dense vector derived from its textual description. Embeddings are generated with cambridgeltl/SapBERT-from-PubMedBERT-fulltext [25], a biomedical language model pre-trained to map semantically equivalent medical concepts to nearby points. For each code, the CLS-token representation of its description is extracted and L2-normalized; pairwise cosine similarities are then converted to cosine distances for the SNF pipeline. Because every code, across all three terminologies, is embedded from its own description into one shared space, distances are directly comparable across terminology types with no per-terminology aggregation. This accommodates the dataset’s partial coverage: not every condition appears in all three systems, yet any pair of codes can still be compared on equal footing.

The clinical view is the only language-dependent component of the framework: SapBERT can be replaced by other biomedical encoder aligned to the target language and terminology. In this case study, we used SapBERT-from-PubMedBERT-fulltext with Brazilian Portuguese descriptions. This encoder is pre-trained primarily on English biomedical text, so applying it to Portuguese is a deliberate approximation whose consequences we examine in the Discussion and quantify in S3 Table.

**Demographic view.** The demographic view represents each diagnosis code as a distribution over the joint sex–age group space. A diagnosis *×* demographic-bin matrix is constructed by summing record counts for each (code, sex, age group) combination, yielding up to twelve bins per code: three sex categories (*Feminino*, *Masculino*, and *Sem Informaçã*) crossed with the four aggregated age groups described above. Pseudocount smoothing (*α* = 0.5) is applied prior to row-normalization to probability distributions, ensuring numerical stability for codes with sparse demographic coverage. Pairwise distances are computed using the chi-square distance [26]. This distance is particularly appropriate here because the demographic signal of interest is the relative distribution of encounters across age and sex strata rather than their absolute volume.

**Temporal view.** The temporal view represents each diagnosis code as a chronological time series indexed by epidemiological week-year pairs, ordered from the earliest to the most recent observation in the dataset. This representation preserves multi-year outbreak dynamics and secular trends. Each series is normalized row-wise to [0, 1] via min-max scaling, mapping the minimum observed count to zero and the maximum to one. This normalization preserves the sparsity structure of the series, keeping weeks with zero cases at zero, while removing magnitude differences between codes with very different absolute volumes.

Pairwise distances are computed using Move-Split-Merge (MSM) [27], which introduces an explicit cost for split and merge operations and has been shown to match or outperform competing measures across a broad range of time series benchmarks [28]. MSM is robust to temporal misalignment and shifted seasonal peaks, a relevant property in a multi-city, multi-year dataset where the timing of epidemic events may differ across geographic units.

#### Syndrome discovery

A candidate syndrome is a group of diagnosis codes that are jointly similar in clinical meaning, demographic profile, and temporal behavior. To recover them, spectral clustering [29] is applied to the fused affinity matrix, partitioning the codes into such groups. The number of clusters *K* is not fixed *a priori* ; a guided sweep over a candidate range (here, 50 to 300) computes two complementary metrics per value: the affinity Z-score and the silhouette score. The affinity Z-score [10] is a permutation-based statistic testing whether the observed cluster structure is more consistent with the fused affinity matrix than expected by chance; it is the primary criterion because it is grounded in the SNF affinity space rather than in Euclidean geometry. The silhouette score is a secondary check on cluster separation.

As a robustness check, the Leiden algorithm [30] is applied to the same fused matrix, sparsified to the 20 nearest neighbors per node, using the Reichardt–Bornholdt configuration model (RBConfigurationVertexPartition) over a resolution sweep from 0.001 to 1.0, where lower resolution yields fewer, larger communities. The final *K* is the smallest value within the plateau of the median affinity Z-score across the ten city-level runs, corroborated by the community counts Leiden produces at comparable granularity. Both criteria and supporting metrics are reported in Results.

Clusters are exported in the Open Syndrome format [14] for direct integration with downstream surveillance systems. One cluster per run is expected to absorb codes lacking a distinctive signal across all three views, identifiable by its disproportionate size and low internal coherence. To assess generalizability, we executed the full pipeline (Fig 1) independently for each municipality. Each city is a self-contained experiment, using the same five input variables and hyper-parameters, with no information shared across runs.

**Fig 1.**
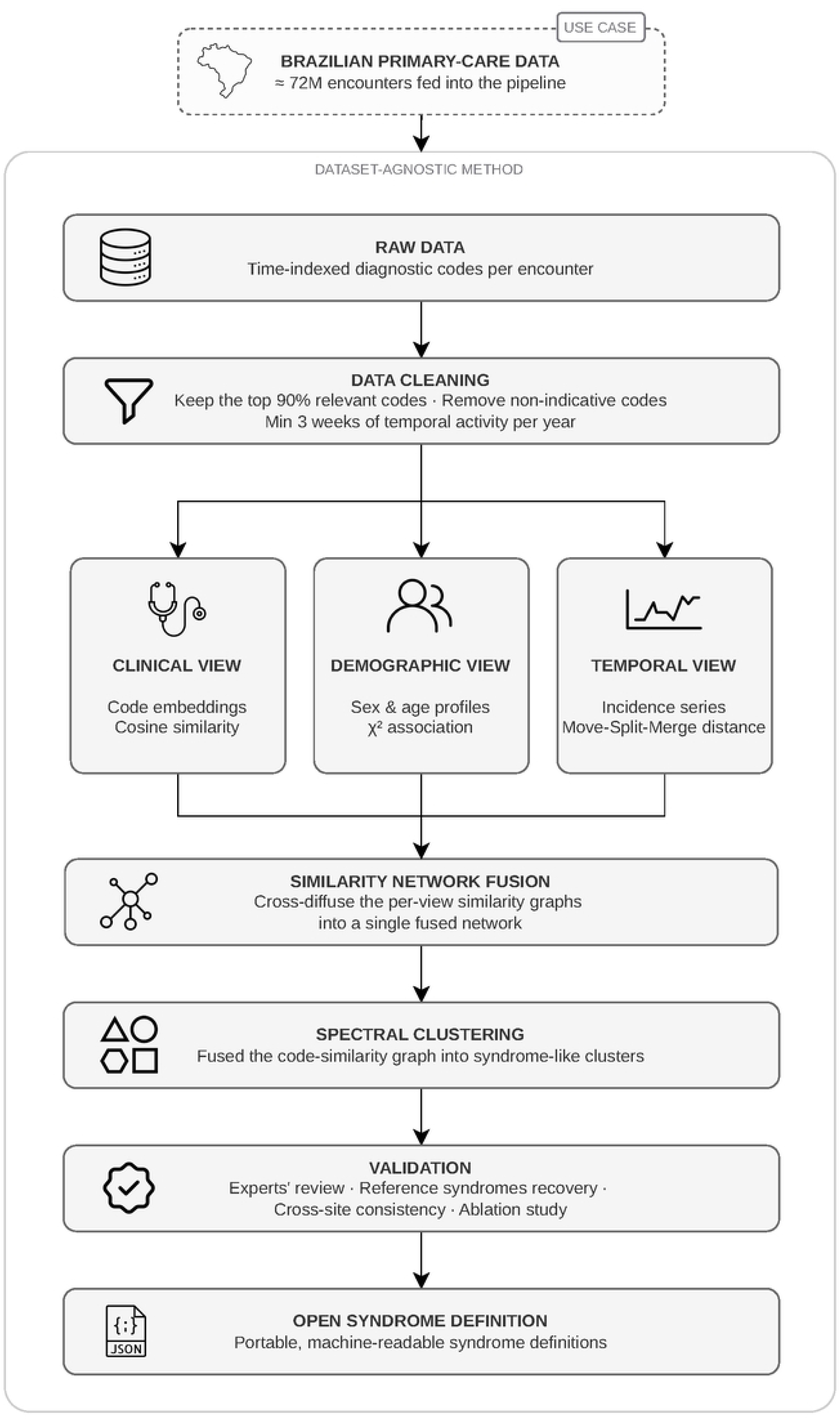
**Overview of the multi-view clustering pipeline for syndromic indicator discovery**. Each diagnosis code is represented across three complementary views (clinical, demographic, temporal), fused via Similarity Network Fusion, and partitioned into syndrome candidates through spectral clustering run independently per city; the resulting clusters are exported as Open Syndrome Definition (OSD) files, while a separate validation branch assesses method performance through ablation, cross-city consistency analysis, and expert review. All codes describing specified signs, symptoms, or diagnoses of diseases or health conditions were labeled as indicative due to their relevance.

**Fig 2.**
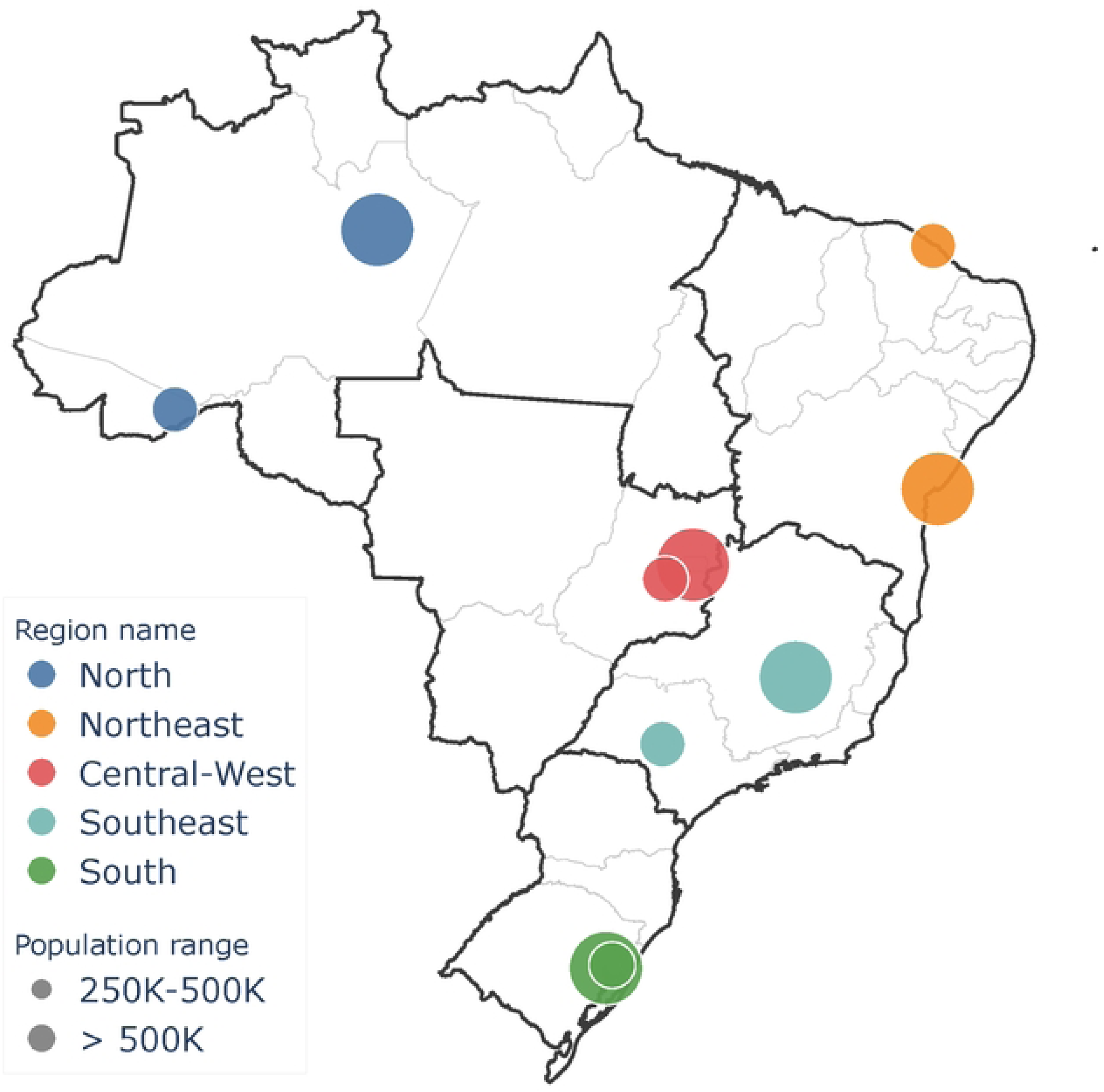
**Geographical distribution of the dataset across Brazilian regions**. Each region is represented by two cities of different population ranges.

### Validation

The validation strategy assesses whether the fused clustering recovers known syndromes without using their definitions as input, whether each view contributes meaningfully to the final partition, and whether the resulting clusters are judged epidemiologically and/or clinically coherent by domain experts.

#### Reference syndrome recovery

The central validation tests whether the framework recovers published syndrome definitions [31] that are not used as input during clustering. Two reference syndromes, arboviruses and ILI, are used, both published and operationally deployed by the Alert-Early System of Outbreaks with Pandemic Potential (ÆSOP) [32] surveillance system in Brazil. They are built from the same data source as the framework, providing an external ground truth directly comparable to the clustering output.

Because PHC diagnoses are typically based on clinical assessment and recorded before laboratory confirmation, both reference syndromes include a broad set of codes to maximize sensitivity. The arbovirus syndrome covers 16 codes for dengue-like arboviral diseases common in Brazil (dengue, Zika, chikungunya); the ILI syndrome covers 50 codes for conditions potentially related to ILI.

Recovery is evaluated with five metrics. *Corpus coverage* is the fraction of reference codes present in the city corpus, the upper bound on recall for that run. *Best-cluster recall* is the fraction of reference codes in the single most concentrated cluster. *Cluster precision* is the fraction of a cluster’s codes that belong to the reference definition; a cluster is *coherent* when its precision is at least *τ* (default 20%). The *number of coherent clusters* counts how many clusters meet this threshold, capturing how far the reference syndrome fragments across the partition. These four metrics are standard in the clustering evaluation literature [33, 34].

A fifth metric, *aggregated recall*, handles a situation that arises naturally here: a reference syndrome may fragment into several coherent sub-clusters. The arbovirus reference, for instance, may split into a dengue/chikungunya cluster and a hemorrhagic-fever cluster, both clinically coherent sub-syndromes rather than artifacts. Because best-cluster recall counts only the single best cluster, it penalizes this split; aggregated recall instead sums the reference codes recovered across all coherent clusters (precision *≥ τ*). A reference syndrome is considered recovered when its aggregated recall reaches 50%. This is a practical adaptation of the BCubed completeness framework [34] to a setting where fragmentation into sub-syndromes is expected and acceptable: BCubed assumes a many-to-one mapping between clusters and classes, whereas here that mapping is the hypothesis under test, not a given. The threshold *τ* is tunable, and sensitivity to its value is reported.

Beyond the quantitative metrics, cluster coherence is assessed qualitatively: for each cluster containing reference codes, the co-grouped non-reference codes are inspected for clinical and/or epidemiological plausibility.

#### Cross-city consistency analysis

Cross-city consistency is evaluated *post hoc* by comparing the cluster partitions produced by independent city-level runs. For each ordered pair of cities (*A, B*) and each cluster *A_i_* in city *A*, the best-matching cluster best match(*A_i_, B*) is the cluster *B_j_* with the highest Jaccard index *|A_i_ ∩ B_j_|/|A_i_ ∪ B_j_|* over their assigned code sets; this maximum value is the match score for the pair (*A_i_, B*).

The consistency threshold *τ* is calibrated using the arboviruses [35] reference syndrome as an anchor, as defined by the ÆSOP [32]. For each pair of cities, the Jaccard score of the cluster most concentrated in arboviruses codes is recorded; *τ* is set to the median of these scores across all city pairs, yielding *τ* = 0.333.

The reference definitions were established by experts and have not been validated city by city, so the data-driven method is expected to reproduce them only partially, and the consistency a reference reaches across cities is itself informative.

A cluster *A_i_* is declared *consistent* if its best-match score is at least *τ* in every other city, the most stringent possible criterion given the study design. Mutual matches, pairs where best match(*A_i_, B*) = *B_j_* and best match(*B_j_, A*) = *A_i_* with both scores *≥ τ*, form the edges of a correspondence graph whose nodes are individual clusters. Connected components spanning all ten cities are treated as cross-city syndrome candidates.

Each component is summarized at two levels of code membership. The *core* set contains codes present in every member cluster of the component, representing the stable diagnostic signal shared across all matched cities. The *union* set contains all codes appearing in at least one member cluster, capturing city-specific extensions to the shared signal.

#### Ablation study

An ablation study quantifies the contribution of each view, and of SNF’s diffusion itself, to the final partition. Eight combinations are evaluated: the full three-view model, three two-view combinations (each removing one view), three single-view models (each using one view alone), and a naive concatenation baseline that horizontally stacks the three affinity matrices and computes pairwise cosine similarity, without the iterative diffusion of SNF.

All conditions share the same distance matrices, affinity construction, and number of clusters as the reference run; only the fusion step changes. For single-view conditions, the affinity matrix serves directly as the fused matrix. For the naive concatenation baseline, no cross-view diffusion is applied, isolating SNF’s non-linear reinforcement from the effect of simply having multiple views.

Four performance metrics are computed per condition. The Adjusted Rand Index (ARI) and Normalised Mutual Information (NMI) measure agreement between each condition’s partition and the full model’s, so a large drop when a view is removed shows that the view substantially shaped the partition. The silhouette score and the affinity Z-score are computed on the fused affinity matrix and measure intrinsic cluster quality independently of the reference partition, indicating whether a change improves or degrades the clustering. The affinity Z-score is the same permutation-based statistic used for cluster-number selection, here applied across ablation conditions with 1,000 permutations, the snfpy [24] default.

#### Expert validation

The candidate syndromes satisfying the cross-city consistency criterion were submitted to a medical doctor with expertise in epidemiological surveillance for blind review. Each candidate was presented as an unordered list of diagnosis codes with their textual descriptions, with no information on the correspondence graph used to assemble the component. Each list was rated on a three-level scale: valid (the codes constitute a coherent syndrome), partially valid (the list is broadly coherent but contains one or more codes whose inclusion is clinically plausible yet not supported by current literature), and not valid (the codes do not form a recognizable syndrome).

## Results

Applied independently to the ten municipalities, the framework recovered the two reference syndromes and adapted them to each city’s local presentation, and surfaced reproducible candidate syndromes in domains not routinely under syndromic surveillance, such as mental health and women’s health. It yielded 200 clusters per city (2,000 total); of these, 7.5% (150) satisfied the pairwise consistency criterion (*τ* = 0.333, *m* = 10), and mutual recognition across all ten cities yielded 47 candidate syndromes, of which 72% (34) were rated fully valid, 9% (4) partially valid, and 19% (9) not valid by a domain expert blinded to the clustering procedure. The number of clusters *K* = 200 was selected at the peak of the median affinity Z-score and corroborated by Leiden community detection (S6 Fig); per-city cluster-size distributions are reported in S2 Table.

### Reference syndrome recovery

Table 4 reports recovery metrics for the two published reference syndromes across all ten cities. The two reference syndromes were recovered with markedly different patterns, reflecting the clinical structure of each.

**Table 4.** Reference syndrome recovery metrics per city. Reference recovery is counted in matched reference categories (ICD-10 codes matched at the 3-character category level, per the ÆSOP convention; CIAP and AB codes matched exactly), while cluster precision is counted in corpus codes. Corpus coverage: fraction of reference codes present in the city corpus. Best-cluster recall (Best-cl. recall): fraction of reference codes in the single most concentrated cluster. Coherent clusters: number of clusters with precision *≥* 0.20. Aggregated recall (Agg. Recall): fraction of matched reference codes across all coherent clusters. Cluster precision: fraction of the dominant cluster’s codes belonging to the reference definition.

| City | Arboviruses |  |  |  |  | ILI |  |  |  |  |
| --- | --- | --- | --- | --- | --- | --- | --- | --- | --- | --- |
|  | Corpus Coverage | Best-cl. Recall | Coherent Clusters | Agg. Recall | Cluster Precision | Corpus Coverage | Best-cl. Recall | Coherent Clusters | Agg. Recall | Cluster Precision |
| Anápolis | 0.33 | 1.00 | 1 | 1.00 | 0.80 | 0.70 | 0.14 | 16 | 0.80 | 0.62 |
| Bauru | 0.22 | 1.00 | 1 | 1.00 | 0.25 | 0.62 | 0.13 | 14 | 0.81 | 0.80 |
| Belo Horizonte | 0.56 | 0.80 | 1 | 0.80 | 0.70 | 0.86 | 0.21 | 14 | 0.79 | 0.64 |
| Caucaia | 0.44 | 0.75 | 2 | 1.00 | 1.00 | 0.78 | 0.13 | 12 | 0.72 | 0.64 |
| Distrito Federal | 0.56 | 1.00 | 1 | 1.00 | 0.86 | 0.90 | 0.22 | 10 | 0.73 | 0.76 |
| Gravataí | 0.22 | 0.50 | 0 | 0.00 | 0.17 | 0.80 | 0.12 | 12 | 0.75 | 0.71 |
| Manaus | 0.56 | 0.80 | 1 | 0.80 | 0.89 | 0.92 | 0.11 | 11 | 0.70 | 1.00 |
| Porto Alegre | 0.56 | 1.00 | 1 | 1.00 | 0.24 | 0.88 | 0.20 | 10 | 0.70 | 0.02 |
| Rio Branco | 0.33 | 1.00 | 1 | 1.00 | 0.57 | 0.82 | 0.12 | 19 | 0.88 | 1.00 |
| Salvador | 0.33 | 1.00 | 1 | 1.00 | 1.00 | 0.86 | 0.14 | 12 | 0.70 | 0.50 |

Arboviruses codes were recovered in nine of the ten cities (Fig 3). In eight of these, a single coherent cluster captured the reference codes (*n*_coherent_ = 1), with best-cluster recall ranging from 0.75 to 1.00 (median 1.00) and aggregated recall equal to best-cluster recall, confirming the absence of fragmentation. Caucaia was the only city where two coherent clusters were identified, with codes distributed across a dengue-dominant and a broader mosquito-borne fever cluster, both clinically coherent (aggregated recall 1.00 vs. best-cluster recall 0.75). Gravatáı did not reach the coherence threshold: with corpus coverage of 0.22, only two arbovirus reference categories were present, and the cluster containing them had precision below 20%, precluding recovery. In the remaining eight cities, aggregated recall ranged from 0.80 to 1.00.

**Fig 3.**
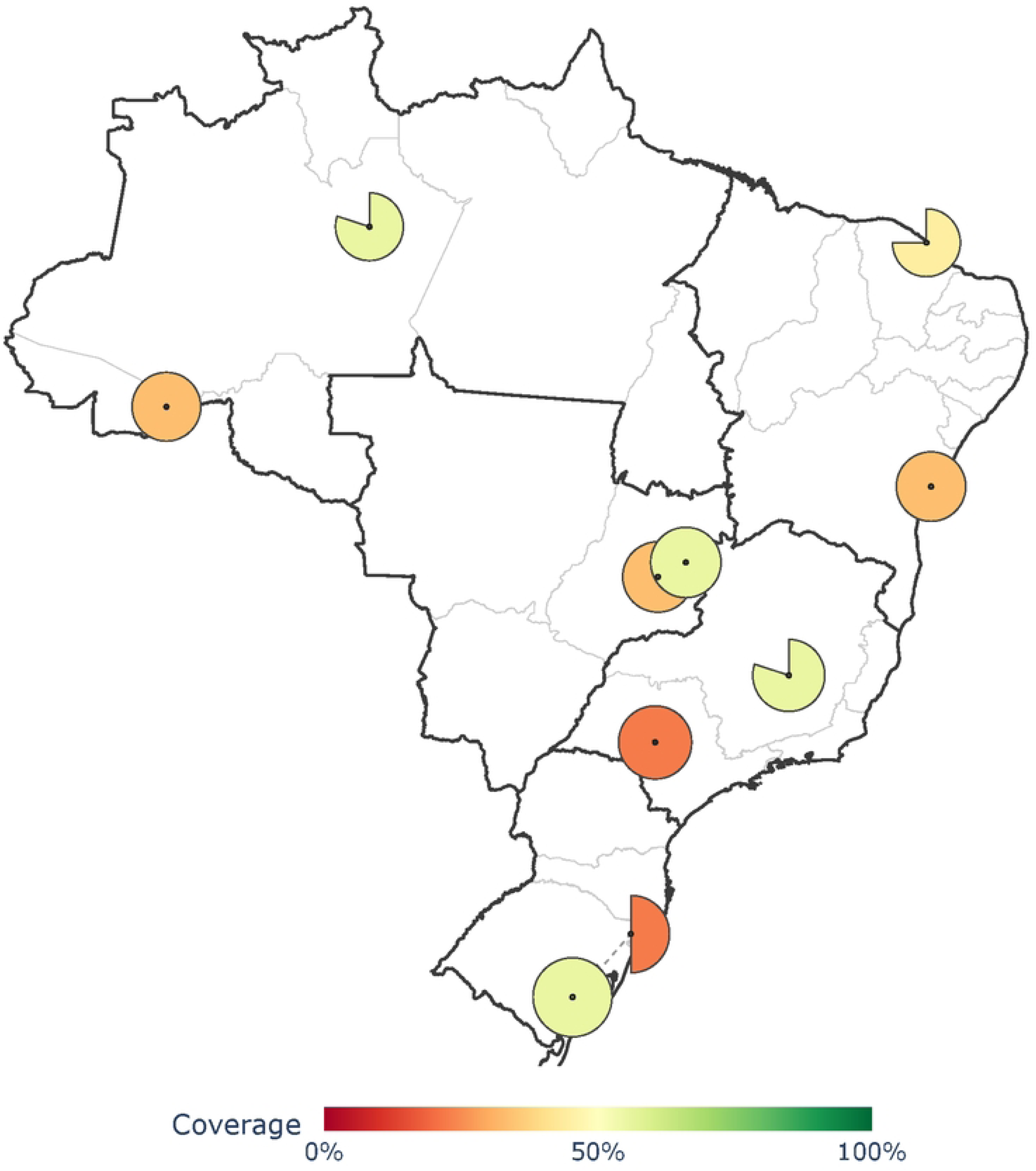
Arbovirus syndrome recovery per city. Each city is shown as a wedge glyph at its geographic location. The filled angular fraction of the glyph encodes best-cluster recall (a full disc indicates a recall of 1.0, a half-disc indicates 0.5). The fill color encodes corpus coverage on a red-to-green scale (0% to 100%), as shown in the legend.

Compared to the arboviruses syndrome, ILI codes exhibited a markedly different recovery pattern (Fig 4). Best-cluster recall was low across all cities (median 0.14), but aggregated recall was substantially higher (median 0.74), recovered across 10 to 19 coherent clusters per city. This fragmentation reflects genuine clinical heterogeneity: influenza, pneumonia, bronchitis, and acute upper respiratory infections have sufficiently distinct demographic profiles and temporal patterns to be separated by the SNF framework into epidemiologically coherent sub-syndromes. Aggregated recall exceeded the 50% recovery threshold in all ten cities (range 0.70 to 0.88). Porto Alegre showed the most extreme dilution, with cluster precision of 0.02 in the dominant cluster, indicating that ILI codes in that corpus are distributed across a large number of clusters without forming a clearly dominant grouping. The number of coherent clusters and their precision varied across cities for both syndromes, so each published definition emerged from a city-specific cluster structure that reflected local coding and epidemiology.

**Fig 4.**
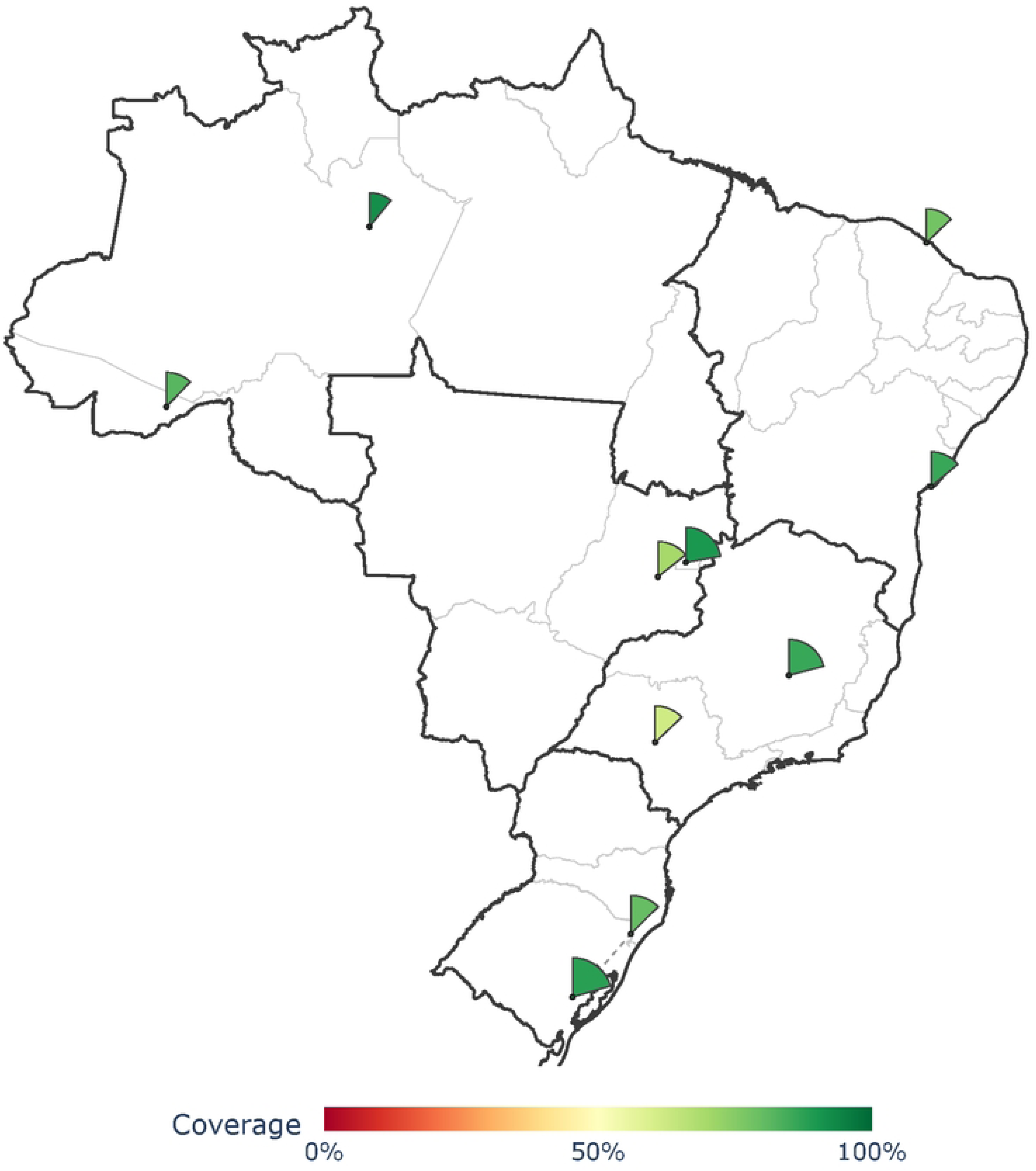
ILI syndrome recovery per city. Glyphs follow the same encoding as in Fig 3: the filled angular fraction of each wedge glyph encodes best-cluster recall and the fill color encodes corpus coverage (red-to-green, 0% to 100%). The narrow wedges reflect the low best-cluster recall obtained for ILI across all cities.

### Expert validation

Inspection of the recovered coherent clusters showed that the additional codes identified outside the reference definition were generally clinically related to the reference codes. For arboviruses, recovery was evident in most cities. In the Distrito Federal, the cluster grouped the six reference codes present in that corpus with a single non-reference code, D69.5 (secondary thrombocytopenia), a clinically meaningful addition given that thrombocytopenia is a recognized complication of dengue [36]; Caucaia and Salvador reached a precision of 1.00 with no non-reference codes. Off-target additions elsewhere were mostly febrile or infectious. The exception was Porto Alegre, whose low-precision cluster absorbed unrelated psychiatric, dermatological, and injury codes, consistent with the dilution expected when a syndrome lacks a clearly dominant grouping.

For ILI, the coherent clusters recovered the influenza and pneumonia families at finer granularity than the three-character reference. The influenza cluster grouped J09, J10, and J11 with their J10.x and J11.x subcodes (influenza with pneumonia, influenza with other respiratory manifestations), and the pneumonia cluster grouped the J12–J18 categories with their subcodes (J12.x, J15.x, J18.x). These subcodes are clinically the same conditions as the reference categories, meaning the framework captures a more specific influenza and pneumonia spectrum than the published definition encodes. A minority of codes lacked clear clinical and/or epidemiological proximity; for instance, A23 (ICPC-2, unspecified risk factor) appeared in a respiratory cluster, attributable to its uninformative description, which failed to anchor the language model embedding to any specific clinical profile.

### Cross-city consistency

Independent runs and consistency analysis across all 10 cities identified 47 candidate syndromes through mutual recognition (Fig 5). Four syndromes are highlighted to illustrate the broad range of syndrome patterns identified: influenza (component 99) and intestinal infections (component 88), both conventional surveillance targets, alongside two conditions that are rarely covered by syndromic surveillance: anxiety (component 9), representing mental health, and irregular or excessive menstruation (component 58), representing women’s health. Most components showed moderate consistency near *τ*, with no city deviating systematically from the others, and the per-city best-match Jaccard scores were strongly concentrated near zero (S5 Fig), indicating that *τ* is non-trivial and that the candidate syndromes form a selective subset of the partition.

**Fig 5.**
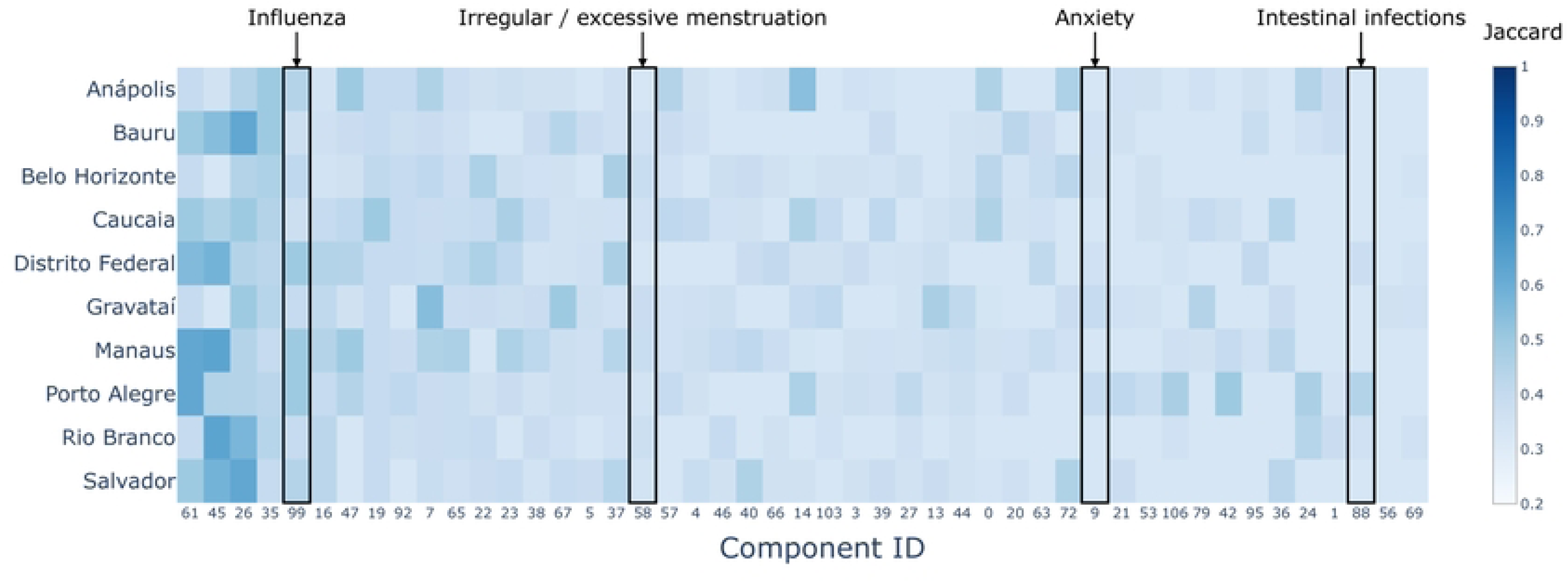
Cross-city consistency of candidate syndromes. Each cell shows the minimum within-component Jaccard between the cluster of that city and the clusters of all other cities in the same component. Components are ordered by decreasing median Jaccard across cities. All values are at or above the arboviruses-calibrated threshold (*τ* = 0.333) by construction. Highlighted components: influenza (99) and intestinal infections (88), both conventional surveillance targets, and anxiety (9) and irregular or excessive menstruation (58), in domains that syndromic surveillance rarely covers.

The proportion of consistent clusters fell monotonically from 100% when a single matching city was required to its all-cities value, with no inflection point (S1 Fig), so the strictest criterion is a deliberately conservative choice.

The expert was blinded to the component structure and assessed each code list independently, rating 72% (34) of the candidate syndromes fully valid and 9% (4) partially valid. Syndromes rated partially valid typically contained one or two codes whose inclusion was epidemiologically plausible but not firmly supported by the literature.

### Ablation study

Fig 6 reports the mean and standard deviation of each performance metric across the ten city-level runs for the eight ablation combinations (results are also available as supplementary material). The full model achieves ARI = 1.00 by construction, as each condition is compared against the full model re-run rather than the stored production run, isolating the contribution of each view from pipeline non-determinism.

**Fig 6.**
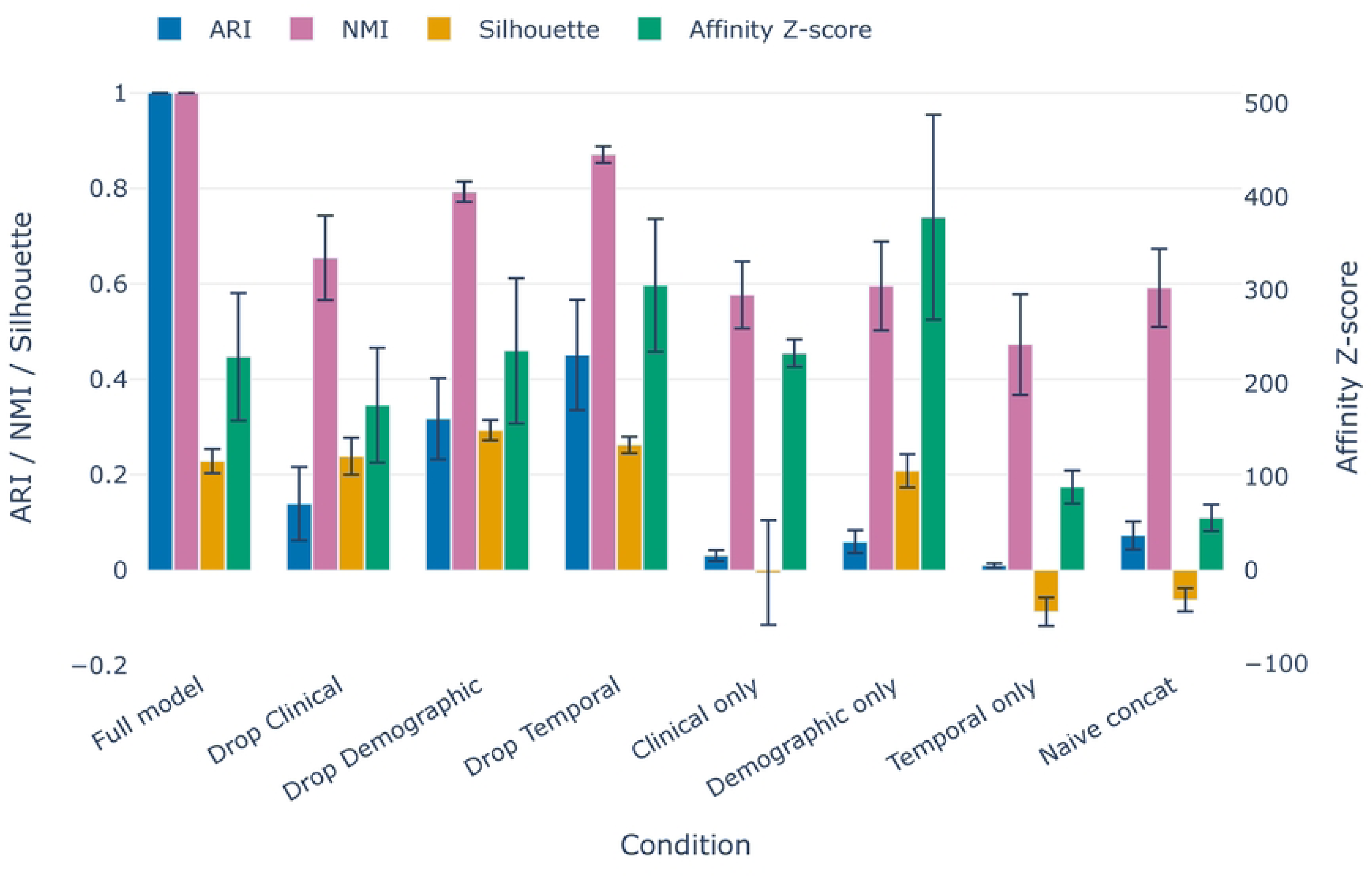
**Ablation study results**. Grouped bar chart of the eight ablation conditions; within each condition, one bar encodes one metric, and color encodes the metric. Whiskers denote *±* one standard deviation across ten independent city-level runs. ARI and NMI measure agreement between each condition and the full-model re-run; silhouette and affinity Z-score measure intrinsic cluster quality in the fused affinity space. ARI, NMI, and silhouette share the left axis ([*−*1, 1]); the affinity Z-score is plotted against the right axis.

The clinical view shaped the partition most: removing it caused the largest decrease in ARI across all cities (0.14 *±* 0.08), indicating that the demographic and temporal views together reproduced little of the full-model partition. In contrast, the temporal view changed the partition least when removed (ARI 0.46 *±* 0.10), and was also the weakest when used alone, producing the lowest affinity Z-score among single views (89.1 *±* 17.5) and a negative silhouette in all cities. Epidemic curves, therefore, do not separate clusters on their own, although removing the temporal view still shifted the fused partition.

The demographic view produced the highest affinity Z-score when used alone (378.2 *±* 109.9), exceeding that of the full model (230.1 *±* 71.4). This reflects the strong internal separation of age and sex profiles in primary care data: conditions concentrated in specific demographic strata form highly cohesive clusters when only the demographic signal is used. By contrast, the full model trades some local cohesion for richer epidemiological structure by incorporating clinical and temporal information that the demographic view cannot capture.

Finally, naive concatenation consistently yielded the lowest affinity Z-score (56.0 *±* 14.3) despite using all three views, so the iterative diffusion of SNF adds structure beyond simply stacking the views.

## Discussion

In this work, we propose an unsupervised multi-view SNF framework for syndrome discovery based on routinely collected health data. The framework recovered both reference syndromes and surfaced candidate syndromes in domains not usually covered by syndromic surveillance, including mental health and women’s health. The recovery of ILI as multiple coherent sub-clusters rather than a single group warrants interpretation beyond a simple recall metric. The SNF framework consistently separated influenza, bacterial pneumonia, bronchitis, and acute upper respiratory infections into distinct partitions across all ten cities, reflecting genuine differences in their demographic profiles and temporal patterns. This fragmentation raises a question that the syndromic surveillance field has not systematically addressed: whether the boundaries of published syndrome definitions reflect epidemiological structure in the data or operational convenience in their construction. In this context, the proposed framework offers a data-driven approach for reassessing existing syndrome definitions to improve overall system performance, a need highlighted by previous work [4].

For instance, the ILI definition used as reference aggregates conditions that share a broad anatomical location and a common route of transmission, but that differ in their age distribution, seasonality, and clinical severity. A method that recovered ILI as a single compact cluster would be capturing the administrative boundary of the definition rather than the structure of the data. The fragmentation observed here suggests that data-driven discovery reveals the internal heterogeneity of expert-curated definitions, and the two together provide a more complete picture of the epidemiological landscape. The aggregated recall metric introduced in this work was designed precisely to accommodate this situation, crediting recovery across coherent sub-syndromes without penalizing clinically meaningful fragmentation.

Additionally, the framework is particularly valuable for capturing local and temporal variability in the data, thereby allowing syndrome definitions to be tailored to the specific context in which they are applied. Because syndrome definitions should be guided by their practical utility [37], this flexibility allows definitions and surveillance priorities to evolve over time while also accommodating previously unforeseen conditions [4, 37]. The diverse range of relevant candidate syndromes identified in our case study illustrates this feature.

Although syndromic surveillance was originally designed to detect infectious disease outbreaks, the scope of public health threats that benefit from early, continuous monitoring extends well beyond communicable diseases. Primary care data captures a broad spectrum of conditions, including chronic non-communicable diseases, acute infections, and conditions that manifest before patients reach hospital settings, making it a complementary surveillance source [35] that has received limited attention in the syndrome discovery literature, which has historically relied on emergency department or hospital admission data. The candidate syndromes validated by the domain expert in this study reflect this breadth: diabetes with complications, intestinal infections, and bacterial pneumonias span both communicable and non-communicable conditions, all managed longitudinally in primary care, and their unsupervised discovery suggests that the epidemiological structure of aggregated primary care data is richer than its administrative origin might suggest.

Among the candidate syndromes, four components grouped codes associated with mental health conditions: anxiety disorders, depressive episodes, bipolar affective disorder, and schizophrenia, all rated epidemiologically coherent by the domain expert. Their emergence from an unsupervised framework operating on routine primary care data reflects the sustained and longitudinal nature of mental health presentations in this setting, which generates the temporal and demographic signal necessary for the clustering to identify them as distinct groups. Nationally defined syndromic surveillance systems rarely include mental health as a monitored domain; the framework recovered these conditions without prior specification, from the same administrative data used for infectious disease reporting.

The cross-city consistency analysis provides the strongest evidence for the generalizability of the framework’s output. Running the pipeline independently on ten municipalities spanning all five Brazilian macro-regions, two population size brackets, and distinct epidemiological profiles, and then identifying which candidate syndromes recur across all runs without any information sharing, transforms consistency into a methodological control rather than a descriptive statistic. A syndrome that emerges independently in Manaus, Porto Alegre, and Ańapolis, cities separated by thousands of kilometers and served by health systems with different funding levels and coding practices, are very unlikely to be attributed to local data artifacts or run-specific hyperparameter choices. The 72% of candidate syndromes rated fully valid by the domain expert, who evaluated each code list without knowledge of the component structure, provides external confirmation that the consistency criterion selects for epidemiological coherence rather than statistical regularity.

Several limitations of this work deserve explicit acknowledgment. Two forms of sparsity are present in this dataset and should be understood as properties of the underlying system rather than deficiencies of the data collection. Code-level sparsity, where many diagnosis codes have low record volume, reflects the structure of medical coding systems: rare conditions, atypical presentations, and underused codes accumulate at the tail of the frequency distribution in any real-world coding dataset. Codes with very low record volume produce unreliable distance estimates [31], which propagate into the affinity matrices and can pull low-frequency codes toward whichever cluster dominates their local neighborhood. Temporal sparsity, where many diagnosis codes have few observations across weeks, reflects genuine epidemiological patterns: conditions that are rare, seasonal, or geographically concentrated will naturally produce sparse time series. The choice of MSM over Dynamic time warping (DTW) for the temporal view and chi-square distance over Euclidean for the demographic view was motivated in part by robustness to these sparsity patterns, but the fundamental constraint remains: distance-based representations require sufficient data volume to be meaningful.

The number of clusters *K* = 200 was held constant across all cities despite substantial variation in the number of active diagnosis codes, ranging from 1,288 in Ańapolis to 3,785 in Distrito Federal. A city-adaptive *K* proportional to the active code set would likely produce more balanced partitions and reduce the size of the residual cluster in larger corpora. Porto Alegre exhibited near-zero cluster precision in the dominant ILI cluster (0.02), with ILI codes distributed across a large number of clusters without forming a clearly dominant grouping, suggesting that local coding practices or data quality issues in that corpus warrant further investigation.

The clinical view inherits the limitations of biomedical language model embeddings applied to administratively generic code descriptions: codes whose descriptions consist primarily of qualifiers such as “unspecified” or “not elsewhere classified” lack the semantic specificity needed to anchor their embedding reliably, and tend to cluster near whichever syndrome dominates their local neighborhood. SapBERT-from-PubMedBERT-fulltext is pre-trained exclusively on English biomedical text, while the code descriptions in this dataset are in Brazilian Portuguese. Biomedical terminology with Latin or Greek roots transfers partially under subword tokenization, but Portuguese-specific clinical vocabulary and grammatical structure are not represented in the pretraining corpus. The ablation study indicates that the clinical view nonetheless contributes uniquely to the final partition (Drop Clinical ARI = 0.14, the largest drop across all conditions), and the SNF diffusion attenuates view-specific noise by reinforcing only cross-view agreement, which mitigates the impact of imperfect embeddings. The recovery of both reference syndromes and the 72% expert validation rate provide empirical evidence that the framework produces coherent clusters under this encoder choice. As a sensitivity check, we re-embedded all code descriptions with two Portuguese-pretrained encoders, BioBERTpt [38] (Portuguese biomedical) and BERTimbau [39] (Portuguese general-domain), and scored the clinical-view geometry against the external ICD-10 chapter hierarchy (S3 Table). The biomedical Portuguese encoder did not significantly outperform SapBERT on most metrics, while the general-domain encoder showed a consistent, significant gain, suggesting language match matters more than domain match here. Both alternatives also shifted generic-code anchoring substantially, a pattern we leave for future characterization. Because the framework is encoder-agnostic and the fused partition is validated end-to-end (72% blind expert validation under SapBERT, with SNF attenuating view-specific noise), the encoder remains a drop-in configuration choice for adopters.

A broader limitation concerns the absence of established benchmarks, public datasets, and evaluation metrics designed specifically for syndrome discovery. Standard clustering metrics such as NMI were not designed for the sparse, high-dimensional affinity spaces produced by SNF, nor for the setting where reference definitions may themselves have low internal epidemiological coherence. The aggregated recall metric introduced here is a practical adaptation, but it is not a substitute for community-agreed evaluation standards. This absence is not accidental: the privacy constraints that motivate privacy-preserving frameworks like the one proposed here also restrict the public release of the data needed to build benchmarks. Establishing evaluation standards for syndrome discovery compatible with aggregated, privacy-compliant data remains an open problem for the community.

Future work can extend the framework in several directions. City-adaptive cluster number selection, using the per-city affinity Z-score peak rather than a fixed *K*, would address the partition imbalance observed in larger corpora. Pre-processing code descriptions to disambiguate generic qualifiers before embedding, for example by appending chapter or category context from the coding system hierarchy, could improve the clinical view for under-specified codes. Active learning represents a natural extension of the validation framework: the expert feedback collected during evaluation, including both valid and partially valid ratings, constitutes a sparse supervisory signal that could guide iterative refinement of the pipeline. Prioritizing clusters with high uncertainty for expert review, and incorporating feedback to adjust representations or thresholds in subsequent runs, would reduce the annotation burden while progressively improving the quality of discovered syndromes. Combining contrastive learning with expert feedback on which code pairs should and should not be considered similar in a surveillance context could produce embeddings that are both clinically relevant and informed by epidemiology, rather than purely semantic, addressing the limitation of generic code descriptions at the embedding level. The threshold calibration mechanism introduced for cross-city consistency, which anchors *τ* to a known epidemiological reference rather than setting it arbitrarily, is a general contribution applicable to any multi-corpus clustering evaluation where a ground-truth anchor exists, and merits formal development as a standalone methodological contribution.

The framework requires only five variables routinely collected by most health system codifying diagnoses: code, sex, age group, epidemiological week, and aggregated encounter count. Its low computational footprint, with no GPU required, and its privacy-preserving design, operating exclusively on pre-aggregated counts rather than individual records, are properties relevant to any setting regardless of income level or regulatory context. Privacy regulations restricting access to granular clinical records exist across all jurisdictions, from the Lei Geral de Proteção de Dados (LGPD) in Brazil to the General Data Protection Regulation (GDPR) in Europe, and aggregated administrative data is frequently the only source available for routine surveillance, even in well-resourced systems. Terminology heterogeneity, another common barrier to cross-system comparability, is addressed by SapBERT’s shared embedding space, which maps semantically equivalent descriptions across ICD-10, ICPC-2, and AB without manual alignment. The clinical view is the only encoder-dependent component of the pipeline: it can be replaced with a biomedical language model aligned to the terminologies and language of the target health system, without altering the demographic or temporal computations.

Each candidate syndrome is exported directly in the OSD [14] format, a structured machine-readable schema designed for interoperability with downstream surveillance systems. This export step can substantially shorten the path from syndrome discovery to operational deployment, addressing one of the core bottlenecks identified in the syndrome definition literature [5].

## Conclusion

We introduce a general framework for syndrome discovery from health records, requiring only five routinely collected variables and three complementary views fused via SNF. A case study on Brazilian primary care data demonstrates that the framework recovers published reference syndromes without their definitions provided as input and identifies additional candidate syndromes that are consistent across ten independent city-level runs spanning Brazil’s five macro-regions. The 72% expert validation rate among cross-city candidates, evaluated blind to the clustering structure, confirms that the consistency criterion captures epidemiological coherence rather than statistical regularity.

The central methodological contribution is twofold: a multi-view general framework for syndrome discovery from medical data, and the use of geographic replication as an evidence standard. This design is applicable beyond Brazil. Any health system that routinely collects aggregated diagnosis counts by demographic group and time period can adopt the same pipeline, replacing the language model with a domain-appropriate alternative where needed. By treating each city as an independent experiment and requiring that candidate syndromes emerge spontaneously across all ten runs, the framework converts consistency into a form of external validation that does not depend on labeled data or predefined definitions.

The limitations identified, fixed cluster number, sensitivity to generic code descriptions, and the absence of field-specific benchmarks point toward concrete directions for future work rather than fundamental constraints of the approach. Syndrome discovery from administrative data is a tool for accelerating and expanding the reach of that knowledge to settings and conditions that manual processes cannot cover at scale.

## Supporting information

**S1 Fig. Sensitivity of cluster consistency to the agreement threshold.** Each point shows the percentage of clusters satisfying the pairwise consistency criterion (*τ* = 0.333) in at least *m* of the ten cities. The dotted line marks the selected threshold *m* = 10, requiring consistency across all municipalities.

**S2 Fig. Effect of the cleaning and preparation steps on the number of unique medical codes (code and code type pairs).** Stages are: Raw (as received); Cleaned (after dropping records with missing year, week or age group); Prepared (after additionally removing codes flagged as non-indicative by the relevant codes reference list and ICD-10 Z-chapter codes). Each bar is annotated with the absolute count and the percentage relative to the Raw stage. The Raw to Cleaned transition removes only 14 records and no codes, indicating very high completeness of the demographic fields, whereas the Cleaned to Prepared transition removes 757 unique codes (6.24%) and about 34.8M records (47.7%), reflecting the dominant contribution of the relevant-code and Z-chapter filters.

**S3 Fig. Per-city data funnel across the cleaning and preparation pipeline.** City-level decomposition of the funnel shown in S2 Fig. For each of the 10 study cities, the three grouped horizontal bars represent the Raw, Cleaned and Prepared stages (light, mid and dark grey, respectively); cities are ordered by their Raw unique-code count. Each bar is annotated with the retention percentage relative to that city’s Raw stage. Cities show consistent retention patterns: the demographic null-drop has negligible impact (Cleaned *≈* 100% of Raw in all sites), while the relevant-codes and Z-chapter filter halves the record volume per city and removes a small but stable fraction of the unique-code repertoire.

**S4 Fig. Indicative classification coverage across clinical terminology systems, by unique code count and record volume**. Stacked bar charts showing the proportion of codes labeled as relevant, non-indicative, or unlabeled (not yet submitted for expert classification) for each of the three terminology systems used in the dataset (ICD-10, ICPC-2, AB). The divergence between ICD-10, ICPC-2, and AB reflects the number of records carrying codes from each terminology.

**S5 Fig. Distribution of best-match Jaccard scores per city, organized by macro-region and population size**. The vertical line marks the arboviruses-calibrated consistency threshold (*τ* = 0.333). The distribution is strongly concentrated near zero in all cities, confirming that *τ* is non-trivial and that consistent clusters represent a selective subset of the partition.

**S6 Fig. Affinity Z-score and Leiden community counts per city.** Affinity Z-score and number of Leiden communities for each of the ten city-level runs. The selected *K* = 200 and resolution *γ* = 0.5 are indicated.

**S1 Table. Expert relevance labeling of the most frequent diagnosis codes.** List of the diagnosis codes accounting for the most frequent 90% of aggregated encounters, each manually labeled by a domain expert. The table reports the code, coding terminology (ICD-10, ICPC-2, or AB), textual description, and assigned label. Codes describing specified signs, symptoms, or diagnoses of diseases or health conditions were labeled relevant; codes for life stages, behavioral factors, healthcare utilization, administrative encounters, or acute trauma and sequelae were labeled non-indicative.

**S2 Table. Cluster size distribution per city (***K* = 200). Size is reported as the number of diagnosis codes. Singletons are clusters of size 1. The residual cluster is the largest cluster in each run. Cluster sizes scale with data volume, and singletons are rare across all cities (0–3 per run).

**S3 Table. Clinical-view embedding sensitivity analysis.** Intrinsic clinical-view geometry under the deployed SapBERT encoder and two Portuguese-pretrained alternatives (BioBERTpt, BERTimbau), scored against external ICD-10 chapters for the full terminology and the Salvador and Belo Horizonte code sets. See S4 Table for per-metric intervals.

**S4 Table. Paired bootstrap for the embedding sensitivity analysis.** Gap of each Portuguese encoder from SapBERT over the full terminology (200 resamples, 95% intervals); an interval excluding zero marks a supported difference.

## Data Availability

Our agreement with the Brazilian Ministry of Health for accessing the referenced databases patently denies authorization of access to any third parties. All requests to access these databases must be addressed to the Brazilian Ministry of Health. https://github.com/anapaulagomes/virola The framework implementation is available at https://github.com/anapaulagomes/virola and archived on Zenodo (DOI: 10.5281/zenodo.XXXXXXX) under the MIT license.

## Acknowledgments

The icons used in this work were created by The Chohans Brand, joalfa, Smashicons, Freepik, iconsax, Anditii Creative, srip, Abdul-Aziz, and Dreamstale, all via Flaticon.

